# Deep learning-based patient stratification for prognostic enrichment of clinical dementia trials

**DOI:** 10.1101/2023.11.25.23299015

**Authors:** Colin Birkenbihl, Johann de Jong, Ilya Yalchyk, Holger Fröhlich, the Alzheimer’s Disease Neuroimaging Initiative

**Author notes:** Corresponding authors: Colin Birkenbihl and Holger Fröhlich. The majority of this work was performed when the author was affiliated with 1 and 2.

## Abstract

Dementia probably due to Alzheimer’s disease (AD) is a progressive condition that manifests in cognitive decline and impairs patients’ daily life. Affected patients show great heterogeneity in their symptomatic progression, which hampers the identification of efficacious treatments in clinical trials. Using artificial intelligence approaches to enable clinical enrichment trials serves a promising avenue to identify treatments.

In this work, we used a deep learning method to cluster the multivariate disease trajectories of 283 early dementia patients along cognitive and functional scores. Two distinct subgroups were identified that separated patients into ‘slow’ and ‘fast’ progressing individuals. These subgroups were externally validated and independently replicated in a dementia cohort comprising 2779 patients. We trained a machine learning model to predict the progression subgroup of a patient from cross-sectional data at their time of dementia diagnosis. The classifier achieved a prediction performance of 0.70 ± 0.01 AUC in external validation.

By emulating a hypothetical clinical trial conducting patient enrichment using the proposed classifier, we estimate its potential to decrease the required sample size. Furthermore, we balance the achieved enrichment of the trial cohort against the accompanied demand for increased patient screening. Our results show that enrichment trials targeting cognitive outcomes offer improved chances of trial success and are more than 13% cheaper compared to conventional clinical trials. The resources saved could be redirected to accelerate drug development and expand the search for remedies for cognitive impairment.

## Introduction

Dementia is a debilitating, progressive condition that is primarily described by cognitive decline. With increasing symptom severity, patients become impaired in their daily life and require full time care, which poses a great burden to patients, caregivers, and society. Seventy five percent of dementia cases are caused by Alzheimer’s disease (AD)^1^. With the recent regulatory approvals of aducanumab^2^, donanemab^3^, and lecanumab^4^ being the only successes in the past 20 years, most clinical trials aiming to identify treatments against cognitive decline in AD have failed^5,6^.

The low success rate of clinical trials aimed at improving cognitive outcomes can be attributed, in part, to the significant variability in how patients’ symptoms progress, even during the earliest stages of the disease^6,7^. This variability poses a statistical challenge and can impede the identification of significant treatment effects. One potential solution to this issue is to increase the sample size of clinical trials to enhance statistical power. However, this approach is costly as it requires more treated patients.

Alternatively, clinical trials can aim for a targeted recruitment of patients that will likely exhibit a faster disease progression and change in cognitive outcomes^8^. This enrichment of patients from a subgroup experiencing a more homogeneous, fast symptomatic progression represents a so-called enrichment trial^9^ and promises several advantages over traditional clinical trials: It can unmask treatments that are only efficacious in specific patient subgroups but fail in the average population^10^, and recruited trial cohorts can be smaller due to the reduced heterogeneity and increase in effect size^11^. The advantages of enrichment trials have also been recognized and promoted by the US Food and Drug Administration (FDA) in 2019. To enable enrichment trials, however, robust patient subgroups with distinct symptom progression patterns must be identified and validated^12^. Furthermore, it must be possible to predict the subgroup membership of an individual from cross-sectional data already available during patient screening. Also, in the light of the recent approvals of three monoclonal antibodies targeting the AD-characteristic amyloid beta pathology^2–4^, the timely prognosis of patients’ likely course of cognitive decline would help to optimize the correct timing and intensity of treatment. Prognostic models designed for this task would also be of immense value from a patient perspective, because they would allow them to better plan their future.

One family of approaches that allows for identifying patient subgroups based on multimodal patient-level cohort data are clustering methods. In the context of AD, such approaches were predominantly applied to cross-sectional data of patients^13–15^. However, cross-sectional clusterings fall short in capturing the longitudinal dynamics of AD dementia, and resulting subgroups can be biased by the disease stages in which patients resided at the time of data collection. Recent studies that take progressive signals into account focussed mainly on exploring the pathology of the disease in the form of imaging biomarkers^16^ rather than the cognitive outcomes directly relevant for clinical trials.

In this work, we apply an artificial intelligence (AI) approach for clustering multivariate clinical disease trajectories of demented AD patients. The resulting subgroups are then externally validated and independently replicated in another cohort study. We construct and validate a machine learning classifier to accurately predict the future progression type of individuals from cross-sectional data only. Finally, we demonstrate the value our classifier could provide to enrichment trials for treatments of cognitive decline by enabling cheaper trials with smaller cohort sizes.

## Materials and methods

### Cohort datasets and patient selection

Two independent cohort datasets were used in this study: the Alzheimer’s Disease Neuroimaging Initiative (ADNI)^17^ and the National Alzheimer’s Coordinating Center (NACC)^18^. Both cohort studies adhered to the declaration of Helsinki and got approval from their institutional review boards. We only included patients that developed dementia probably due to AD during the runtime of their respective cohort study. Further, patients must have had at least one follow-up assessment after their dementia diagnosis and visits prior to it were excluded. This led to 283 analyzable patients for ADNI and 2779 for NACC, with a median of two years follow-up respectively. Three years after diagnosis, 70 patients were available in ADNI and 1080 patients in NACC.

### Multivariate patient trajectory clustering

To cluster patients into symptom progression subgroups, we used our previously published VaDER approach that was specifically designed with longitudinal clinical data in mind^19^. VaDER is a deep learning approach that clusters multivariate time-series data and imputes missing values implicitly during model training. Hyperparameters were optimized following the procedure described in de Jong et al.^19^: We evaluated several possible models using different configurations of hyperparameters (including the number of sought after subgroups) and selected the hyperparameters of the best performing model. Model performance was measured by comparing the prediction strength of the clustering induced by the trained model against a random clustering of the same data^20^. To determine the optimal number of clusters, we selected the smallest number that showed a significant difference from random clustering **(Figure S1)**. Selected hyperparameters are presented in **Table S1**.

We clustered patients based on their trajectories of three major clinical scores measuring symptom progression: the Mini Mental State Examination (MMSE), Clinical Dementia Rating Sub of Boxes (CDRSB), and Functional Activities Questionnaire (FAQ). Patient trajectories were aligned on their dementia diagnosis to avoid biases introduced through patients being in different clinical disease stages at their study baseline. Considering the average length of currently ongoing phase 3 trials for cognitive treatments enrolling early AD patients (approximately 24 months, **Table S2**) and the longitudinal follow-up of ADNI and NACC, we clustered trajectories spanning up to 3 years. Each clustering was repeated 40 times and the final subgroup assignments of patients were based on a consensus clustering across the repeats.

### Cluster validation and replication in external data

To evaluate the robustness and validity of our identified patient subgroups, we conducted an external validation of ADNI-derived subgroups in NACC and, additionally, performed a replication of the analysis starting with NACC data. For the external validation, we applied the ADNI-trained clustering model to NACC to determine whether resulting subgroups of NACC patients resembled those identified in ADNI. Furthermore, we aimed to replicate the results by starting with a new, independent clustering of NACC to see if we would get an optimal clustering that was similar to the one achieved under the ADNI-trained model. We further externally validated this NACC-derived clustering in ADNI. Finally, we assessed the concordance between patient assignments within each dataset under both clustering models.

### Building a machine learning classifier for cluster prediction

We built machine learning classifiers aiming to predict the progression cluster membership of individuals based on their cross-sectional data at the time of dementia diagnosis. For this, we used the XGBoost algorithm which is based on an ensemble of decision trees and can handle missing values^21^. The classifiers were trained and evaluated in a 10 times repeated nested 8-fold cross-validation with hyperparameter optimization in the inner 8-fold cross-validation and model evaluation in the outer one (details in the Supplementary Material).

As more data modalities were available in ADNI than in NACC^22^, we built two separate classifiers: a multimodal classifier based on the ADNI data, and another classifier using only features common between ADNI and NACC (in the latter referred to as ‘common predictor’ classifier). The ‘common predictor’ version of the classifier allowed for an external validation.

The multimodal classifier incorporated demographic information (7 features), clinical assessments and their subscores (11 assessments amounting to 66 features in total), biomarkers (62 magnetic resonance imaging (MRI) derived brain region volumes, 3 positron emission tomography, 4 cerebrospinal fluid), and genetic variables (APOE ε4 status, 75 disease pathway perturbation scores; see Supplementary Material for details on pathway score calculation and the Supplementary Spreadsheet for a list of all predictors). Individual MMSE questions were summed into subscores as described in the Supplementary Material. Due to the increased requirements on the available data compared to the initial clustering, the sample size of ADNI reduced to 230 patients for this analysis.

For the ‘common predictor’ version of the classifier the number of available features decreased to 28, now comprising only APOE ε4 status, clinical, and demographic features (1, 22, and 5 features, respectively). The clinical features are the Trail Making B score, Montreal Cognitive Assessment score, Digit Span score, and summary scores and subscores of the MMSE, Clinical Dementia Rating, and FAQ. A detailed list is provided in the Supplementary Spreadsheet. This classifier was trained on the larger NACC dataset and externally validated on ADNI.

### Simulating the impact of patient enrichment on clinical trial design

We estimated a potential reduction in trial cohort sample size enabled through an enrichment of patients with ‘fast’ symptom progression while maintaining adequate statistical power. This analysis was performed by applying the NACC-trained ‘common predictor’ classifier to ADNI to mirror a scenario with classifier-independent data. Stratified patient recruitment was simulated by only including patients into our hypothetical trial cohort whose predicted probability of belonging to the ‘fast’ progressing cluster exceeded a threshold. The specifications of the hypothetical trial were adapted from recent clinical trials for early to mild AD dementia **(Table S2)**: As a primary outcome, we used the change from baseline in CDRSB since dementia diagnosis and considered a trial runtime of up to 24 months. The treatment arm was simulated using an effect size of 27%, a value that was observed for the recently approved lecanemab^24^. Effect size was calculated as Cohen’s d. Conservatively, we did not simulate the effect size dependent on the progression rate, but uniform over all patients. Effects were only emulated in patients who actually experienced a worsening of the outcome during the 24 months. Outcomes for patient’s showing improvement or no change remained unaltered. The theoretical control arm consisted of the same patients without simulated treatment effects. For a power analysis, we considered a two-sided t-test. Following the lecanemab trial, the required statistical power was considered 90% at an alpha level (type I error) of 0.05^4^.

We approximated the impact of reducing the trial sample size through patient enrichment in terms of adverse events and monetary expenses using the ‘Clarity AD’ trial for lecanemab as guidance^4^. We ignored patient drop-out in our estimations. Annual treatment costs of lecanemab amount to $26,500 per patient^25^. As exact information about the costs of patient screening in the lecanemab trial was missing, we assumed the same costs that were previously estimated for the aducanumab trial with $6957 per screened patient^26^.

Adverse events were simulated based on their frequencies of occurrence observed in the original lecanemab trial^4^. Amyloid-related imaging abnormality (ARIA) diagnosis and monitoring incurrences were assumed to involve an additional physician visit ($128) and monthly MRIs ($353 per scan)^26^ for a mean duration of 4 months until resolvement^4^.

## Results

### Identification, validation, and replication of two AD dementia progression subtypes

When clustering the ADNI patients’ trajectories, we identified two distinct symptom progression subgroups that separated patients into ‘slow’, and ‘fast’ progressors **(Figure 1A, Figure S1A)**. One 177 of the 283 patients (63%) were assigned to the ‘fast’ progressing cluster, 106 (37%) to the ‘slow’ progressors. Over the 36 month period, ‘fast’ progressing patients experienced symptom worsening of 6.38 (95% CI: [5.32, 7.45]) for CDRSB, −9.24 (95% CI: [−7.23, −11.25]) for MMSE, and 13.19 (95% CI: [11.22, 15.17]) for FAQ. In contrast, on average, ‘slow’ progressing patients showed significantly reduced worsening with 1.85 (95% CI: [1.30, 2.40]), 1.83 (95% CI: [0.87, 2.78]), and 5.59 (95% CI: [4.17, 7.01]), for CDRSB, MMSE, and FAQ, respectively.

**Figure 1:**
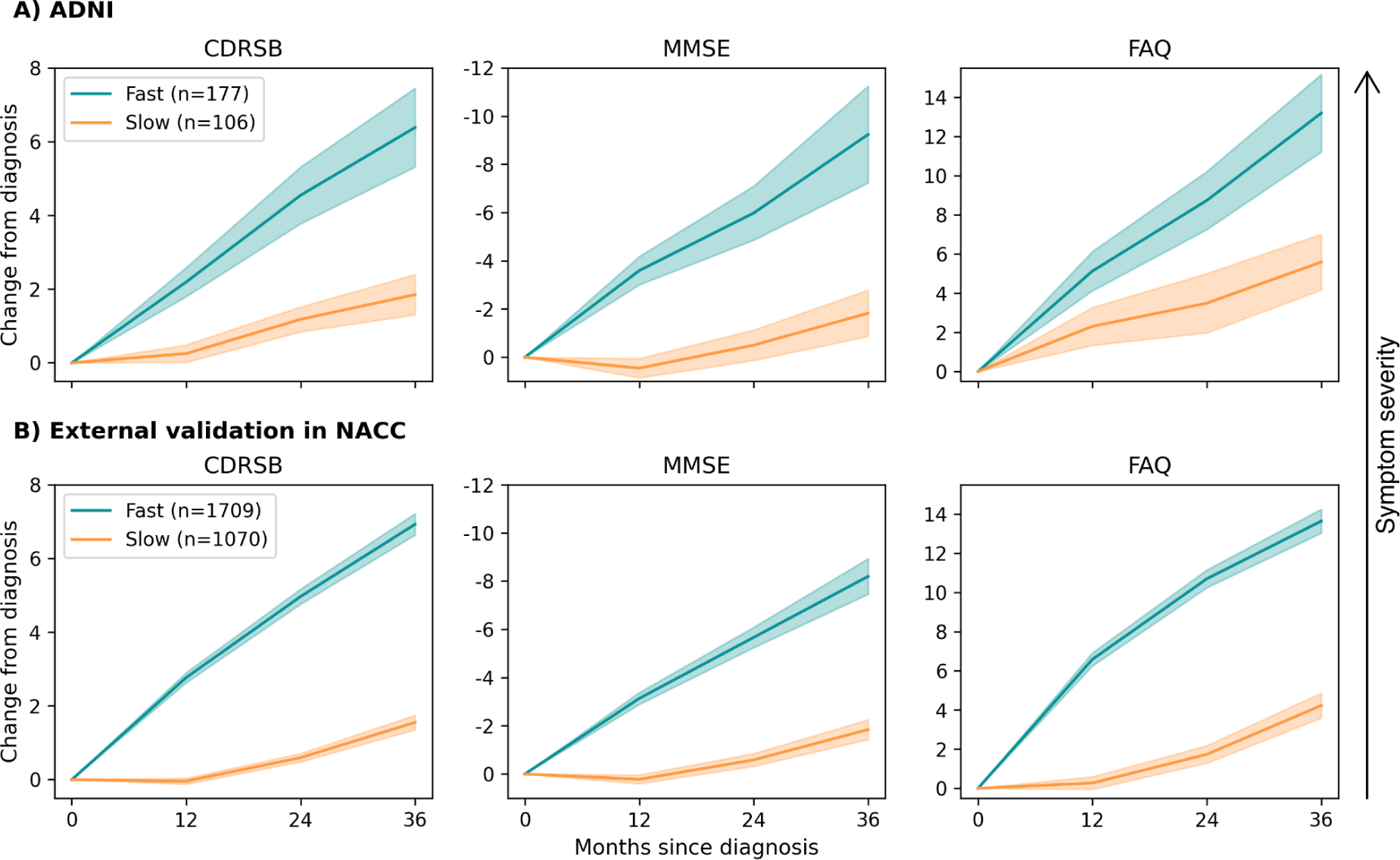
Average symptom progression trajectories of subgroups identified in ADNI and NACC under the ADNI-trained clustering model. For each clinical assessment, the severity of the symptom increases along the y-axis from bottom to top.

When externally validating the clustering achieved in ADNI by applying the ADNI-trained model to patient trajectories from NACC, we obtained two subgroups of NACC patients that were highly similar to those identified in ADNI **(Figure 1B)**. Matching the proportions in ADNI closely, about 61% (1709) of the NACC patients exhibited a ‘faster’ progression, while 39% (1070) experienced ‘slower’ symptom progression. Also the observed empirical average trajectories of NACC subgroups were similar to those identified in ADNI. On average, the ‘fast’ progressors showed symptom worsening of 6.93 (95% CI: [6.63, 7.22]) for CDRSB, −8.20 (95% CI: [−7.46, −8.94]) for MMSE, and 13.64 (95% CI: [13.04, 14.25]) for FAQ over 36 months. ‘Slow’ progressing patients symptoms increased by 1.55 (95% CI: [1.35, 1.75]), −1.84 (95% CI: [−1.43, −2.25]), and 4.22 (95% CI: [3.59, 4.86]), for CDRSB, MMSE, and FAQ, respectively.

Beyond externally validating the ADNI clustering in NACC, we investigated whether the clustering of NACC under the ADNI-trained model would be concordant with an independent clustering achieved by training a new model on NACC. Indeed, a two-subgroup partition provided the best clustering solution for NACC, again splitting patients into ‘fast’ and ‘slow’ progressors **(Figure S1B)**. Comparing the subgroup assignment of NACC patients into ‘fast’ or ‘slow’ progressors under the NACC-trained model and ADNI-trained model showed an agreement of 88%. We additionally applied the NACC-trained model on ADNI for external validation. Again, a highly similar clustering was found with 82% of the ADNI patients assigned to the same subgroup using the independent ADNI-trained and NACC-trained models, respectively.

### Characterization of symptom progression subgroups

We compared demographic variables (age, education, biological sex) between ‘slow’ and ‘fast’-progressors and found no statistically or clinically significant differences at the time of dementia diagnosis in ADNI **(Table 1)**. In NACC, a statistically significant difference was identified for patient age, however, of insignificant clinical relevance (1.01 years, 95% CI: [0.31, 1.70]). Further, in ADNI, we observed a statistically significant but small difference at patient’s dementia diagnosis for the CDRSB score (0.49, 95% CI: [0.05, 0.94]). In NACC, FAQ and MMSE scores at diagnosis differed statistically significantly across subgroups (−1.11 [−1.49, −0.73]; and 1.86 [0.99, 2.72], respectively), but once again with small effect sizes. A further significant difference was found in the distribution of APOE ε4 carriers across NACC subgroups, with 9.33% [13.1%, 5.53%] more ε4 carriers being assigned to the ‘fast’ progressing group.

**Table 1:** Summary statistics describing the empirical distribution of clinical and demographic variables in symptom progression subtypes identified in ADNI and NACC at time of patient’s dementia diagnosis. Numerical variables: mean ± standard deviation; the difference between subtypes is quantified as the difference in means and its 95% confidence interval. Categorical variables: Proportion of APOE e4 carriers and female patients, respectively; differences across subtypes are quantified as the difference in proportions and its 95% confidence interval.

| | N | MMSE | FAQ | CDRSB | Age<br>(Years) | Education<br>(Years) | APOE ε4<br>positive (%) | Female<br>(%) |
| --- | --- | --- | --- | --- | --- | --- | --- | --- |
| ADNI Clusters |  |  |  |  |  |  |  |  |
| **Slow** | 106 | 24.21 ± 2.71 | 11.46 ± 5.9 | 3.98 ± 1.36 | 75.32 ± 6.95 | 15.91 ± 2.73 | 67.92 | 25.47 |
| **Fast** | 177 | 23.55 ± 3.33 | 12.79 ± 5.62 | 4.48 ± 1.81 | 75.53 ± 7.04 | 15.9 ± 2.88 | 76.84 | 36.16 |
| **Difference [95% CI]** |  | -0.66 [-1.5, 0.17] | 1.33 [-0.22, 2.88] | **0.49 [0.05, 0.94]** | 0.2 [-1.68, 2.09] | 0.0 [-0.76, 0.75] | -8.91 [-19.84, 1.65] | 10.69 [-0.58, 21.04] |
| **NACC Clusters (predicted using the ADNI-trained model)** |  |  |  |  |  |  |  |  |
| **Slow** | 1078 | 25.02 ± 4.12 | 14.57 ± 12.44 | 3.75 ± 3.19 | 77.68 ± 9.12 | 15.64 ± 3.22 | 48.42 | 51.58 |
| **Fast** | 1716 | 23.91 ± 4.2 | 16.43 ± 10.31 | 3.88 ± 2.29 | 78.68 ± 9.18 | 15.64 ± 3.14 | 57.75 | 52.39 |
| **Difference [95% CI]** |  | **-1.11 [-1.49, -0.73]** | **1.86 [0.99, 2.72]** | 0.14 [-0.07, 0.34] | **1.01 [0.31, 1.70]** | 0.0 [-0.24, 0.24] | **-9.33 [-13.1, -5.53]** | 0.81 [-2.99, 4.61] |

Comparison of cerebrospinal fluid biomarkers of AD pathology, namely amyloid beta 42, phosphorylated tau, and total tau, did not reveal any significant differences between the two clusters in neither ADNI or NACC (Mann-Whitney U-test p > 0.05 for all biomarkers in both cohorts). Also regarding amyloid PET, no significant difference was identified (p > 0.05).

### Predicting symptom progression subtype from cross-sectional data at time of diagnosis

The multimodal machine learning classifier trained on ADNI was able to differentiate between ‘slow’ and ‘fast’ progressing patients with an average area under the receiver operating characteristic (AUC) of 0.69 ± 0.02 estimated via a 15 times repeated cross-validation **(Figure 2)** and an area under the precision-recall curve (AUC-PR) of 0.60 ± 0.03.

**Figure 2:**
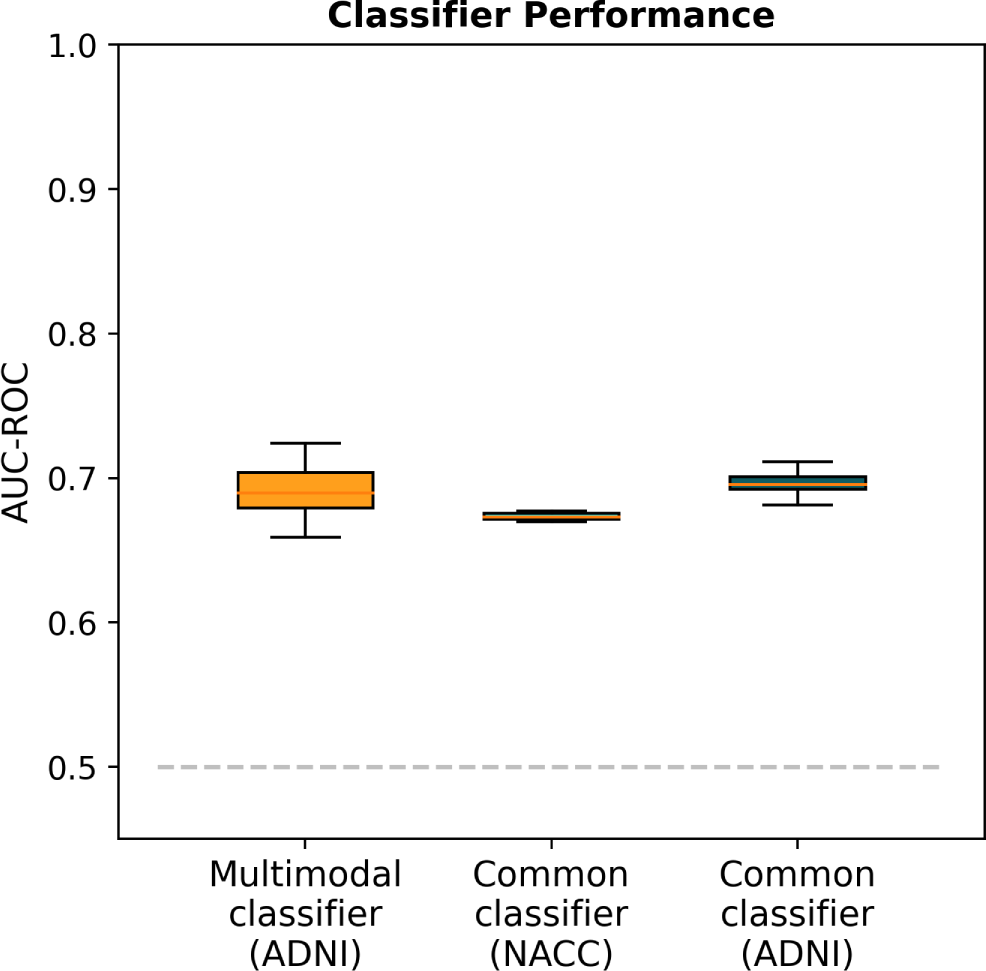
Performance of machine learning classifiers differentiating between ‘fast’ and ‘slow’ progressors at dementia diagnosis, averaged across 15 repeats. The dataset on which the respective performance was evaluated is shown in parenthesis on the x-axis. The application of the common classifier to ADNI represents an external validation.

To externally validate our classifier, we developed a second version that only incorporated features present in both ADNI and NACC. Since NACC holds a substantially larger sample size, we used this dataset to train and internally validate the classifier and used ADNI for external validation. In internal validation on NACC, the classifier achieved 0.67 ± 0.003 AUC **(Figure 2)** and an AUC-PR of 0.58 ± 0.003. External validation in ADNI demonstrated a performance of 0.70 ± 0.01 AUC and 0.56 ± 0.01 AUC-PR, which was similar to both the multimodal classifier’s performance in ADNI and the model’s internal validation scores on NACC **(Figure 2)**, and thereby indicated the model’s generalizability. Feature importance is shown in Figure S3.

### AI-based stratification to enrich cohorts with specific symptom progression subtypes

We emulated an enrichment of a hypothetical clinical trial cohort with patients experiencing ‘fast’ symptom progression by applying our ‘common predictor’ classifier to ADNI. The predicted probability for each patient to belong to the ‘fast’ progressing subgroup was used as an exclusion criterion and patients with predictions below a selected threshold were excluded from the trial.

Without stratification, at their time of dementia diagnosis, 144 of the 230 (62.6%) analyzed ADNI patients belonged to the ‘fast’ progressing subgroup. Expectedly, increasing the classifier threshold required for patient inclusion caused a decrease in the number of patients remaining in the hypothetical trial cohort **(Figure 3B)**. Simultaneously, however, the proportion of ‘fast’-progressors among the remaining patients rose consistently **(Figure 3A)**. After stratifying ADNI using the classifier at a threshold of 0.65, 51.7% of patients (95% CI: [49.9%, 53.6%]) remained in the cohort. The resulting stratified cohort contained 73.4% ‘fast’ progressors (95% CI: [67.5, 79.2]).

**Figure 3:**
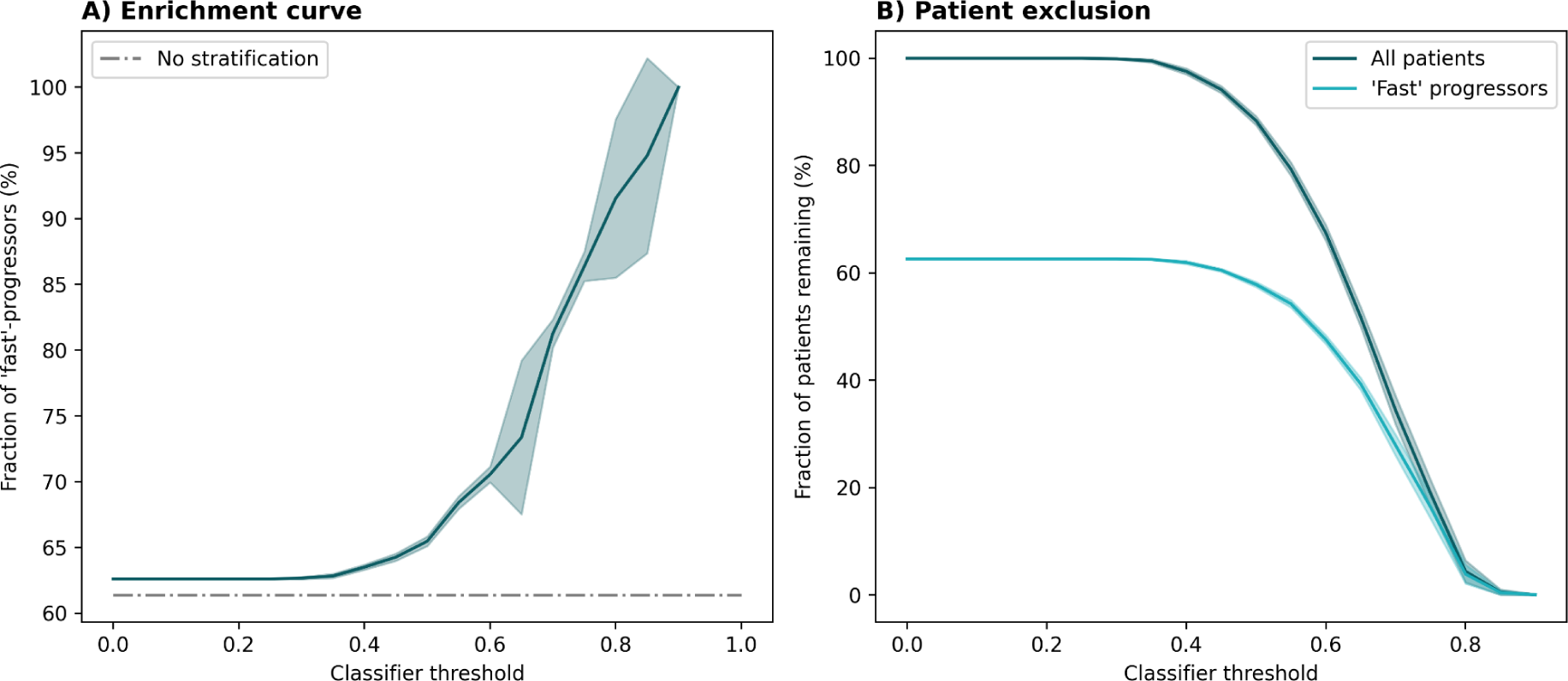
The impact of patient enrichment using ADNI as a hypothetical trial cohort. Mean trajectories and CIs were calculated across 15 repeats, each with a newly trained model. **A)** Enrichment of ‘fast’ progressors with higher classifier thresholds. **B)** Decrease in sample size with higher classifier thresholds.

### Reducing the required trial cohort sample size through patient enrichment

To estimate a possible reduction in trial cohort sample size achieved through patient enrichment with our ‘common predictor’ classifier, we performed a statistical power analysis. The parameters for this analysis were taken from recent trials with cognitive endpoints targeting early AD **(Table S2)**, primarily the ‘Clarity AD’ trial evaluating lecanemab which identified an effect size of 27%^4^.

As previously discussed, increasing the required prediction threshold for patient inclusion led to a more homogeneous, faster progressing trial cohort on average **(Fig. 3A)**. This can lead to measuring greater effect sizes which opens the opportunity to reduce the cohort sample size while maintaining appropriate statistical power (here, 90%). The relationship between the classifier prediction required for patient enrollment and the resulting potential for sample size reduction is presented in **Figure 4**. Assuming a threshold of 0.65, for example, the classifier enabled a sample size reduction of 36.8% (CI: [34.0%, 39.5%]).

**Figure 4:**
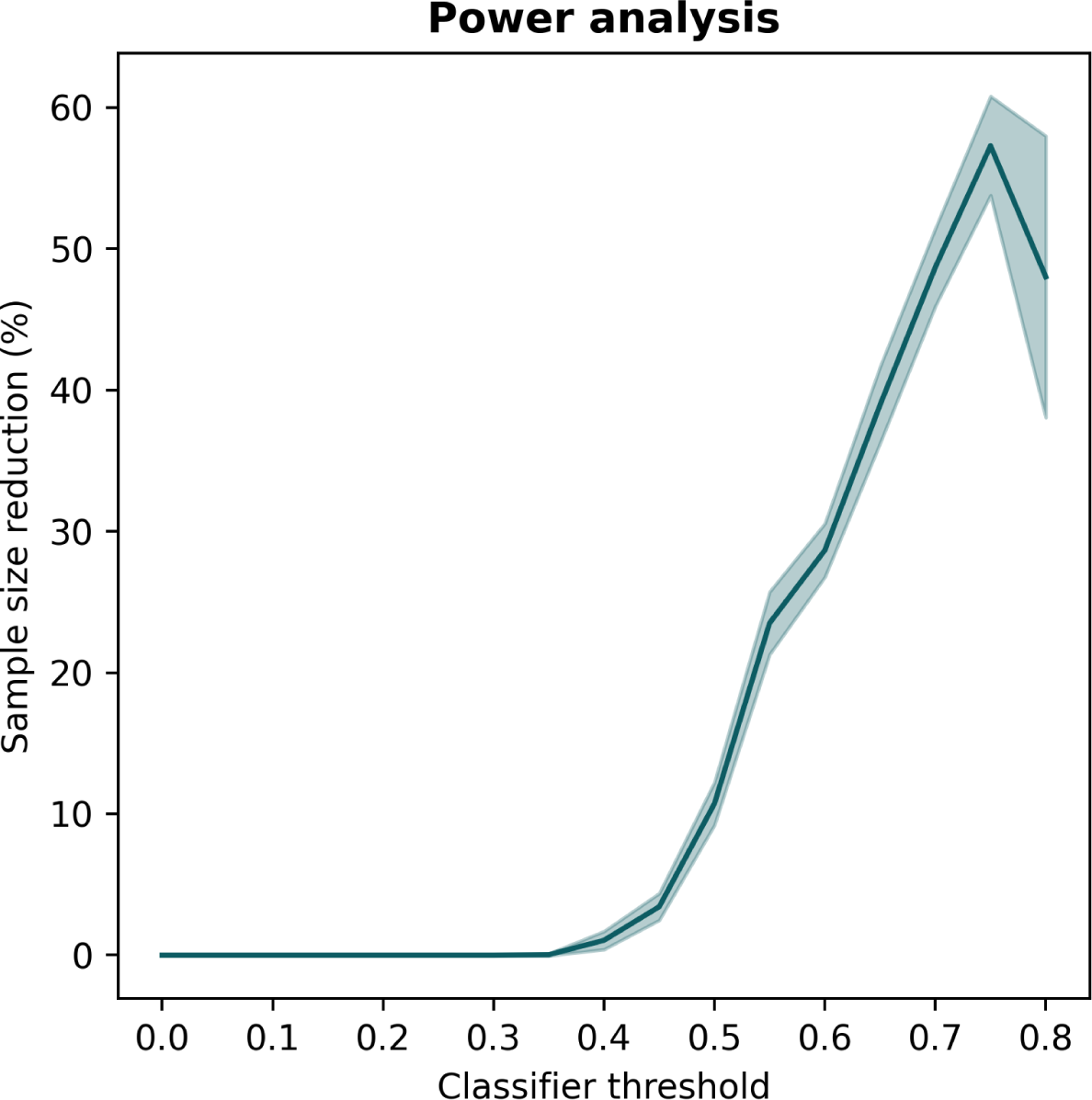
Possible reduction in trial cohort size while maintaining statistical power at 90% in relation to classifier threshold employed for patient enrichment. The line displays the mean trajectory calculated across 15 repeats. The shade represents the 95% CI. Larger confidence intervals at higher thresholds are due to lower abundance of individuals with higher scores.

### Estimating the impact of patient enrichment on economical expenses and patient harm

We approximated the economical impact of patient enrichment with our proposed classifier on trials by counterbalancing the possible sample size reduction with the additional expenses of increased patient screening **(Table 2)**. We assumed a hypothetical clinical trial similar to the successful ‘Clarity AD’ trial for lecanemab^4^ with 24 months runtime and CDRSB as primary outcome. The externally validated ‘common predictor’ classifier was used for patient enrichment. As a classifier prediction threshold for patient recruitment 0.65 was selected.

In ‘Clarity AD’, 1795 patients were enrolled from 5967 screened individuals^4^. Applying our classifier during patient recruitment could reduce the sample size by 36.8% (661 patients) while maintaining 90% statistical power **(Fig. 4)**. This would enable an enrichment trial of the same power by enrolling 1135 participants. Recruiting the enriched trial cohort would require screening 867 additional patients, increasing the trial costs by $6,031,719. The 24 months treatment costs for the lecanemab group (50% of the cohort) would amount to approximately $47,594,000 for the conventional trial and $30,077,500 for the enrichment trial.

During the original lecanemab trial, 593 participants experienced adverse events^4^ while only 375 patients would be affected in an enrichment trial (218 patients reduction). In the enrichment trial, we would further assume 83 fewer serious adverse events than in the conventional trial (144 versus 227). With respect to ARIA, 102 less cases would occur in an enrichment trial, reducing the monitoring costs for ARIA by $157,080 (from $428,120 to $271,040).

The estimated expenses for a conventional lecanemab trial sum up to approximately $89,534,539 while the enrichment trial would cost $77,892,678, thus saving 13% ($11,641,861) of the total costs. Notably, this estimate represents a lower bound neglecting the expenses for treating heterogeneous adverse events and longitudinal monitoring procedures, such as regular neuroimaging.

## Discussion

In this work, we utilized deep learning to identify two distinct AD dementia patient subgroups exhibiting ‘slow’ and ‘fast’ symptom progression, respectively. The subgroups were robustly discovered in two independent datasets and externally validated. Using a machine learning classifier, we were able to predict the longitudinal progression subtype of an individual patient with good performance relying only on cross-sectional data collected at the time of their dementia diagnosis. By emulating a clinical trial employing prognostic patient enrichment using this classifier, we demonstrated the statistical, economical, and patient health related benefits enrichment trials hold over conventional clinical trials in clinical AD dementia.

Instead of relying solely on cross-sectional data, as is commonly done for clustering AD dementia patients^13–15^, we utilized a longitudinal approach that clusters the multivariate progression of patient trajectories^19^. For clustering variables, we deliberately focussed on clinical outcomes of high relevance to clinical trials. Such outcomes could be biased by differences in the pathological disease stage of patients. However, we could not find any significant differences in key biomarkers of AD pathology between the two subgroups.

### Enriching clinical trial cohorts with specific symptom progression subtypes

We developed two classifiers that predicted the cognitive symptom progression subtype of an individual with good prediction performance. We could not observe a significant improvement in performance when using the proposed multimodal classifier over the feature-reduced ‘common predictor’ classifier. The reasons for this could lie in the relatively limited number of patients for which the additional biomarker measurements were available and the high complexity of this data which could warrant greater sample sizes to fully exploit using machine learning approaches. In theory, however, additional predictors could improve the sensitivity and specificity of a classifier. Especially for borderline cases, the continuous nature of biomarker values could allow for more nuanced predictions compared to the mainly categorical features used in the ‘common predictor’ classifier.

Our developed machine learning classifiers were not designed for decision support in a clinical care setting and we do not believe that their performance is sufficient for this task. Instead, we deliberately aimed at building classifiers for patient screening in clinical enrichment trials. In such a setting, the patient enrichment using a machine learning classifier gains its power through an application across a substantial number of potential trial participants. While any classifier that performs above chance-level will lead to an enrichment of a sought-after patient subgroup given that enough patients are screened, better performing classifiers are more cost effective as fewer patients need to be screened to achieve the required cohort size and homogeneity. Similar conditions apply to the threshold placed on classifier predictions, which represents an arbitrary decision that weights the expanse of patient screening against the achieved homogeneity of the resulting trial cohort.

Here, we focussed on prognostic enrichment, however, another promising route would be predictive enrichment based on disease subtypes^27^. The discrimination of patients on a mechanistic-level could enable novel clinical trial designs such as umbrella trials^28^ for AD, but would not guarantee an increased homogeneity in the outcome of interest.

### Implications of enrichment trials targeting cognitive decline

Our results indicate that the inter-patient heterogeneity in cognitive symptom progression could hamper clinical trials especially when their duration is shorter than two years. During this period, the identified ‘slow’ progression subgroup experienced only minute cognitive decline and even highly efficacious treatments would show small effect sizes. Previously, the solution to this statistical challenge was often considered to involve increasing sample sizes^29,30^, however, we argue that enrichment trials present a promising alternative with additional benefits.

In our simulated hypothetical enrichment trial, we found that an enrichment trial aiming at a cognitive outcome could be conducted with significantly smaller cohort sizes as compared to currently ongoing and previously successful trials. Simultaneously, this would lead to cheaper clinical trials and less participant harm caused.

### Limitations

Our presented approximations of monetary expenses in the context of conventional clinical trials and enrichment trials are limited in many ways and the final amounts represent only rough estimates. Additional costs, such as follow-up care and treatments for non-ARIA side-effects have been neglected, and no additional expenses for applying the classifier and accordingly prolonged screening phases were considered.

## Data availability

The used datasets are publicly available at https://adni.loni.usc.edu/ and https://naccdata.org/.

## Acknowledgments

Data collection and sharing for this project was funded by the Alzheimer’s Disease Neuroimaging Initiative (ADNI) (National Institutes of Health Grant U01 AG024904) and DOD ADNI (Department of Defense award number W81XWH-12-2-0012). ADNI is funded by the National Institute on Aging, the National Institute of Biomedical Imaging and Bioengineering, and through generous contributions from the following: AbbVie, Alzheimer’s Association; Alzheimer’s Drug Discovery Foundation; Araclon Biotech; BioClinica, Inc.; Biogen; Bristol-Myers Squibb Company; CereSpir, Inc.; Cogstate; Eisai Inc.; Elan Pharmaceuticals, Inc.; Eli Lilly and Company; EuroImmun; F. Hoffmann-La Roche Ltd and its affiliated company Genentech, Inc.; Fujirebio; GE Healthcare; IXICO Ltd.; Janssen Alzheimer Immunotherapy Research & Development, LLC.; Johnson & Johnson Pharmaceutical Research & Development LLC.; Lumosity; Lundbeck; Merck & Co., Inc.; Meso Scale Diagnostics, LLC.; NeuroRx Research; Neurotrack Technologies; Novartis Pharmaceuticals Corporation; Pfizer Inc.; Piramal Imaging; Servier; Takeda Pharmaceutical Company; and Transition Therapeutics. The Canadian Institutes of Health Research is providing funds to support ADNI clinical sites in Canada. Private sector contributions are facilitated by the Foundation for the National Institutes of Health (www.fnih.org). The grantee organization is the Northern California Institute for Research and Education, and the study is coordinated by the Alzheimer’s Therapeutic Research Institute at the University of Southern California. ADNI data are disseminated by the Laboratory for Neuro Imaging at the University of Southern California.

The NACC database is funded by NIA/NIH Grant U01 AG016976. NACC data are contributed by the NIA-funded ADCs: P30 AG019610 (PI Eric Reiman, MD), P30 AG013846 (PI Neil Kowall, MD), P30 AG062428-01 (PI James Leverenz, MD) P50 AG008702 (PI Scott Small, MD), P50 AG025688 (PI Allan Levey, MD, PhD), P50 AG047266 (PI Todd Golde, MD, PhD), P30 AG010133 (PI Andrew Saykin, PsyD), P50 AG005146 (PI Marilyn Albert, PhD), P30 AG062421-01 (PI Bradley Hyman, MD, PhD), P30 AG062422-01 (PI Ronald Petersen, MD, PhD), P50 AG005138 (PI Mary Sano, PhD), P30 AG008051 (PI Thomas Wisniewski, MD), P30 AG013854 (PI Robert Vassar, PhD), P30 AG008017 (PI Jeffrey Kaye, MD), P30 AG010161 (PI David Bennett, MD), P50 AG047366 (PI Victor Henderson, MD, MS), P30 AG010129 (PI Charles DeCarli, MD), P50 AG016573 (PI Frank LaFerla, PhD), P30 AG062429-01(PI James Brewer, MD, PhD), P50 AG023501 (PI Bruce Miller, MD), P30 AG035982 (PI Russell Swerdlow, MD), P30 AG028383 (PI Linda Van Eldik, PhD), P30 AG053760 (PI Henry Paulson, MD, PhD), P30 AG010124 (PI John Trojanowski, MD, PhD), P50 AG005133 (PI Oscar Lopez, MD), P50 AG005142 (PI Helena Chui, MD), P30 AG012300 (PI Roger Rosenberg, MD), P30 AG049638 (PI Suzanne Craft, PhD), P50 AG005136 (PI Thomas Grabowski, MD), P30 AG062715-01 (PI Sanjay Asthana, MD, FRCP), P50 AG005681 (PI John Morris, MD), P50 AG047270 (PI Stephen Strittmatter, MD, PhD).

## Author Contributions

Designed and supervised the project: HF; analyzed the data and implemented algorithms: CB, IY, JDJ; drafted the manuscript: CB, HF, JDJ; all authors have read, revised, and approved the manuscript.

