## Supplementary Material for "Deep learning-based patient stratification for prognostic enrichment of clinical dementia trials"

Corresponding authors: Colin Birkenbihl (cbirkenbihl a t mgh.harvard.edu) and Holger Fröhlich (holger.froehlich a t scai.fraunhofer.de)

### *Hyperparameter optimization for VaDER-based trajectory clustering*

| Training data | Hidden nodes per layer | Learning rate | Batch size | Alpha |
|---|---|---|---|---|
| **ADNI** | [32, 1] | 0.0001 | 16 | 1 |
| **NACC** | [32, 8] | 0.00001 | 16 | 1 |

**Table S1:** Final hyperparameters of the VADER models trained on ADNI and NACC, respectively. Alpha: Weighting parameter between the KL divergence and reconstruction loss.

[1] Alzheimer's Disease Neuroimaging Initiative: Data used in the preparation of this article were obtained from the Alzheimer's Disease Neuroimaging Initiative (ADNI) database (adni.loni.usc.edu). As such, the investigators within the ADNI contributed to the design and implementation of ADNI and/or provided data but did not participate in analysis or writing of this report. A complete listing of ADNI investigators can be found at http://adni.loni. usc.edu/wp-content/uploads/how_to_apply/ ADNI_Acknowledgement_List.pdf

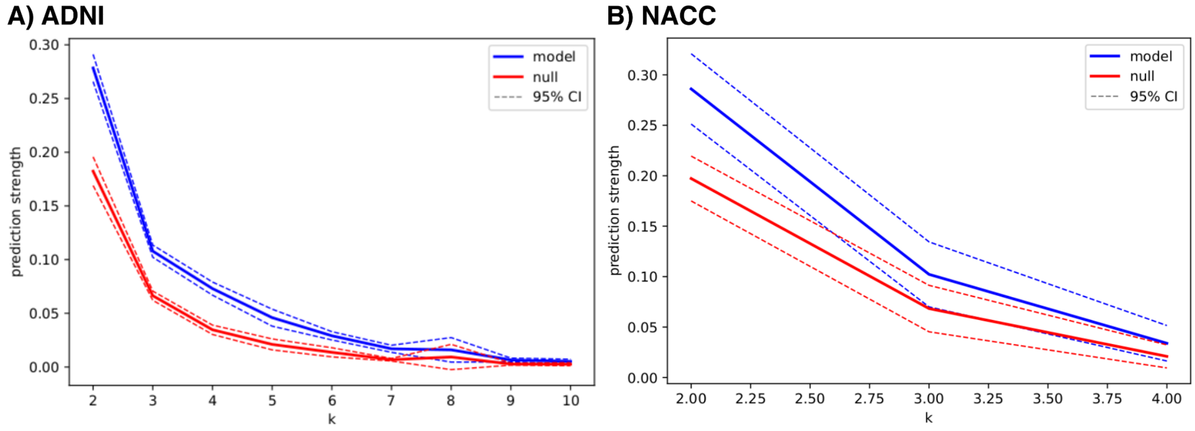


**Figure S1:** Prediction strength of (A) the ADNI-trained clustering mode and (B) the NACC-trained clustering model compared to a random clustering (null) for different numbers of sought after clusters (k).

#### *MMSE subscore calculation*

Subscores for the MMSE were calculated by summing questions that belonged to a common topic. Individual questions are abbreviated according to the ADNI data naming convention.

- MMSE Time: MMDATE, MMYEAR, MMMONTH, MMDAY, MMSEASON
- MMSE Place: MMHOSPIT, MMFLOOR, MMCITY, MMAREA, MMSTATE
- MMSE Registration: MMBALL, MMFLAG, MMTREE
- MMSE Attention: MMD, MML, MMR, MMO, MMW
- MMSE Recall: MMBALLDL, MMFLAGDL, MMTREEDL
- MMSE Language: MMWATCH, MMPENCIL
- MMSE Repetition: MMREPEAT
- MMSE Commands: MMHAND, MMFOLD, MMONFLR, MMREAD, MMWRITE, MMDRAW

#### *Calculating pathway perturbation scores*

We initially applied DisGeNET[1] to retrieve 640 single nucleotide polymorphisms (SNPs) that were putatively associated with Alzheimer's Disease (AD). Only manually curated associations mentioned in at least one publication were used. We expanded this set by all those, which were in strong linkage disequilibrium ( $r^2 > 0.8$) and mapped LD blocks to closest genes via Haploreg v4.1[2]. In addition, we considered a mapping of SNPs to genes via analysis of cis expression quantitative trait loci (cis-eQTLs). More specifically, we used GTex[3] to obtain a list of significant (false discovery rate < 5%) expression quantitative trait loci (eQTLs) in different brain regions. Altogether, LD block analysis plus eQTLs resulted in a list of 22,438 SNPs. Note that both approaches can result in a mapping of one SNP to several genes.

We considered a mapping of these genes to AD-specific pathways defined in NeuroMMSig[4]. For each pathway, a perturbation score was calculated by counting the unique number of non-reference alleles occurring within the body of genes associated with that pathway, and this number was divided by the total number of SNPs mapping to that pathway.

#### *Results*

| **Treatment** | **Run time** | **N** | **Endpoint** | **NCT ID** |
|---|---|---|---|---|
| ALZ-801 | 78 weeks | 300 | ADAS-cog 13 | NCT04770220 |
| Caffeine | 30 weeks | 248 | NTB scores | NCT04570085 |
| Donepezil | 26 weeks | 240 | MMSE | NCT04661280 |
| Lecanemab | 18 months<br>Extension phase until | 1906 | CDR-SB | NCT03887455 |

| | | | | |
|---|---|---|---|---|
| | month 69 | | | |
| Aducanumab | 78 weeks | 1653 | CDR-SB | NCT02477800 |
| Semaglutide | 104 weeks | 1840 | CDRSB | NCT04777396 |
| Gantenerumab | 116 weeks | 984 | CDRSB | NCT03444870 |
| Donanemab | 76 weeks | 1800 | iADRS | NCT04437511 |
| TRx0237 | 52 weeks | 598 | ADAS-cog 11 | NCT03446001 |
| Guanfacine | 12 weeks | 160 | ADAS-Cog | NCT03116126 |

**Table S2:** Recent phase 3 trials enrolling early to mild AD patients taken from clinicaltrials.gov on the 16th February 2023.

#### *Developing the XGBoost classifiers*

Our XGBoost classifiers were built using the xgboost python framework[5]. Hyperparameters were optimized using Bayesian optimization[6]. The objective function of the hyperparameter optimization was the area under the precision-recall curve. The number of base learners (i.e., boosting steps) was determined using early stopping. Hyperparameters to be optimized were:

- "gamma": uniform distribution [0.1, 6.0]
- "learning_rate": log uniform distribution [1e-4, 2]
- "max_depth": integer uniform distribution [3,12]
- "subsample": discrete uniform distribution [0.6, 1] with a step size of 0.1

### Results

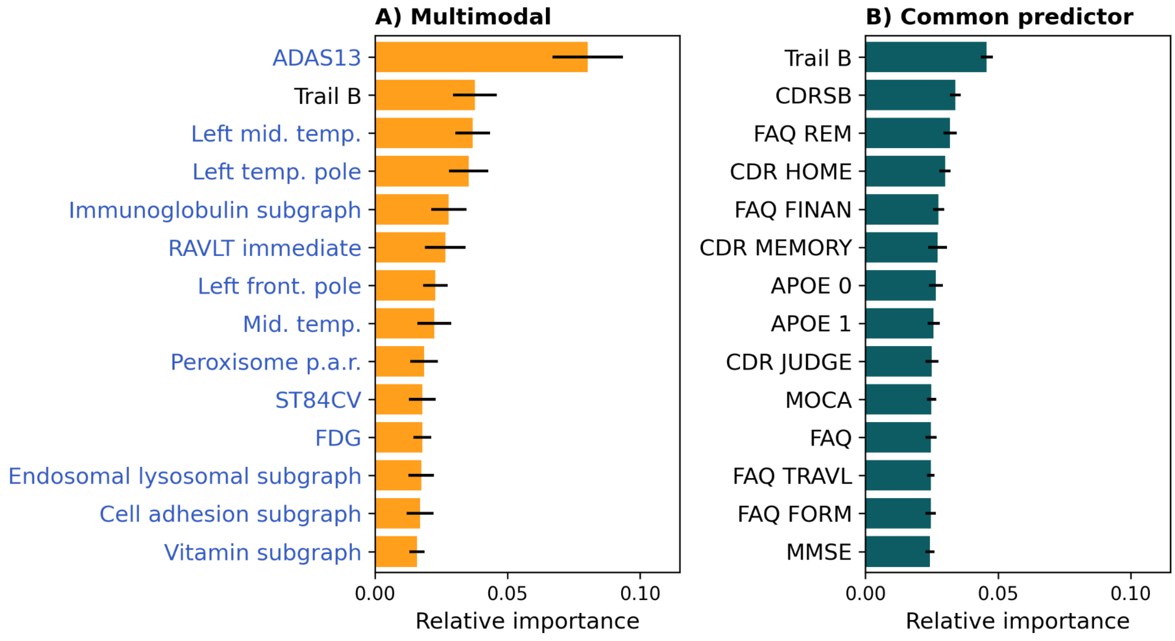


**Figure S3:** Importance of top 15 predictors used in our classifiers. Importance is measured as the predictor gain averaged across all base learners of the XGBoost model. **A)** the multimodal classifier trained and cross-validated on ADNI. Blue font marks features that were exclusively available in ADNI. Peroxisome p.a.r.: Peroxisome proliferator activated receptor subgraph. **B)** the NACC-trained classifier using only predictors that were available in both NACC and ADNI.
